# *APOE* ε4 Effects on Cognitive Decline and Progression to Dementia are Sequentially Mediated through Plasma P-tau217 and Hippocampal Atrophy in an Asian Cohort

**DOI:** 10.64898/2026.11.22.26368713

**Authors:** Cheuk Ni Kan, Joyce Ruifen Chong, Daniel Casado-Rodrigo, Yuek Ling Chai, Narayanaswamy Venketasubramanian, Boon Yeow Tan, Juan Helen Zhou, Christopher Li-Hsian Chen, Mitchell K. P. Lai, Chin Hong Tan

## Abstract

**Background:** The *APOE* ε4 allele is a major genetic risk factor for Alzheimer’s disease (AD) but underlying pathophysiological mechanisms may vary across populations. In Asian cohorts, the relative contributions of plasma phosphorylated tau (p-tau217), neurodegeneration, and cerebrovascular disease to ε4-associated cognitive decline remain unclear.

**Objective:** We investigated biomarker pathways linking *APOE* ε4 to cognitive trajectories and incident dementia.

**Methods:** *APOE* genotyping, plasma p-tau217 measurement, and neuroimaging were conducted at baseline in the Singapore Harmonization longitudinal cohort. Participants underwent annual comprehensive neuropsychological assessments for five years. We examined ε4 group differences in p-tau217, regional subcortical volumes, cortical thickness, and white matter hyperintensities (WMH). Serial mediation analyses examined mediating effects of plasma p-tau217 and neuroimaging markers on ε4 associations with cognitive decline and dementia risk.

**Results:** *APOE* ε4 was present in 29.4% of cohort (N=659; age=73.5±8.0 years; female=56.4%; Chinese ethnicity=83.6%). Carriers had higher p-tau217, reduced entorhinal thickness, and smaller hippocampus, amygdala, accumbens, and putamen volumes, but no WMHdifferences. Pathways from *APOE* ε4 through p-tau217 to hippocampal volume (*β*=- 0.009, 95% CI=-0.015, −0.004, 13.8%) and entorhinal thickness (*β*=-0.008, 95% CI=- 0.014, −0.004, 13%) significantly mediated steeper global and domain-specific cognitive decline. *APOE* ε4 was associated with increased dementia risk (HR=2.35, 95% CI=1.37-4.06), partially mediated by p-tau217 and hippocampal volume (9%). Direct ε4 effects were minimal and WMH did not influence these associations.

**Conclusions:** *APOE* ε4 contributes to cognitive decline and incident dementia through an AD-specific cascade captured by increased p-tau217 and hippocampal atrophy, providing population-specific insights into *APOE* ε4-related pathways in older Asian adults.

## Introduction

Alzheimer’s disease (AD) is an etiologically complex neurodegenerative disorder influenced by a myriad of genetic and physiological factors. Among the known genetic factors (1), the ε4 allele of the *APOE* gene, encoding apolipoprotein E4, is recognized as a major contributor to elevated lifetime risk and earlier age of onset for late-onset sporadic AD (2). Of the multiple mechanistic pathways implicated (3), *APOE* ε4 is thought to influence the pathogenic cascade at multiple points, including increased amyloid-β aggregation, impaired amyloid-β clearance leading to the accumulation of neuritic plaques and tangles, and subsequent tau-mediated neurodegeneration (4, 5).

Building on these mechanistic insights into the role of *APOE* ε4 in AD, it remains crucial to determine whether these processes exert similar effects across diverse populations and ethnicities. While much has been established on the pathophysiological relationship between *APOE* ε4 and AD risk (6), prior evidence was predominantly from cohorts of European descent and the extent of the neurobiological and cognitive impact of *APOE*ε4 remains unclear in non-European populations. Among major ancestral groups, East Asians exhibit one of the lowest frequencies of *APOE* ε4 (7), yet may lack other protective genetic variants and carry ethnic-specific promoter variants that increase AD susceptibility (8). These distinctions in *APOE* polymorphisms may in turn influence how *APOE* ε4 affects brain dysfunction and cognitive decline in Asian populations.

Comparative neuroimaging studies have shown that associations between *APOE* ε4 and markers of cerebral aging, such as reduced cortical thickness and brain volumes, were more pronounced in East Asian than in European-ancestry cohorts (9, 10).

Clinically, the effects of *APOE* ε4 on cognitive function in European populations were influenced by both cerebral small vessel disease and AD-related pathology, including amyloid-β, phosphorylated tau (p-tau), and medial temporal atrophy (11, 12). By contrast, studies in Asian cohorts (13, 14) have found little evidence of cerebrovascular disease (CeVD) in ε4-associated cognitive impairment, pointing instead to a greater role of AD pathology.

As emphasized in the revised framework for AD diagnosis and staging (15), precision plasma p-tau biomarkers can serve as sensitive indicators of underlying AD neuropathologic changes. Recent findings in East Asian populations with concomitant CeVD show that plasma p-tau at threonine 217 (p-tau217) accurately identifies cerebral amyloid-β positivity (16, 17), although the contribution of *APOE* ε4 to cognitive function in relation to plasma p-tau and cerebral changes has not been systematically examined. Together, these observations underscore the need for population-specific studies to elucidate the *APOE* ε4-related pathways to clinical outcomes.

To address key knowledge gaps on whether *APOE* ε4 effects are attributable to AD- specific mechanisms or CeVD in Asian populations, we hypothesized that 1) *APOE* ε4 carriers would have higher plasma p-tau217 and reduced brain volumes in AD-related regions; 2) *APOE* ε4 would be associated with faster cognitive decline and higher dementia risk, replicating clinical outcomes in this understudied cohort; and 3) these clinical effects of *APOE* ε4 would be mediated sequentially through plasma p-tau217 and neurodegeneration markers, independent of CeVD.

## Methods

### Study cohort

The HARMONIZATION longitudinal cohort comprises 700 participants who were enrolled between August 2010 and October 2020 from the National University Hospital, St. Luke’s Hospital, and the community in Singapore. Details on the recruitment criteria and study design of the cohort have been extensively described (18). In brief, all participants underwent extensive clinical examination, blood test, and comprehensive neuropsychological assessment annually, and neuroimaging biennially for up to 5 years. Baseline cognitive status was diagnosed using information from the clinical evaluation, a locally validated comprehensive neuropsychological test battery (19), and neuroimaging assessment.

Alzheimer’s disease dementia was diagnosed according to the National Institute of Neurologic and Communicative Disorders and Stroke – Alzheimer’s Disease and Related Disorders Association criteria (20). Vascular dementia was diagnosed according to the National Institute of Neurologic Disorders and Stroke – Association Internationale pour la Recherche et l’ Enseignement en Neurosciences criteria (21). Participants with ≥ 1 impaired domain on the neuropsychological test battery but without functional loss, hence not meeting criteria for dementia, were diagnosed with cognitive- impairment no-dementia (CIND). Participants without any impaired cognitive domains were diagnosed with no cognitive impairment (NCI).

This study was conducted in accordance with the Declaration of Helsinki and was approved by the National Healthcare Group Domain-Specific Review Board (2018/01098), with written informed consent obtained before enrollment.

### Neuropsychological assessment

Cognitive performance was assessed annually using a comprehensive neuropsychological test battery that has been validated in the cohort (22). The battery assessed cognitive domains of visual and verbal memory (Hopkins Verbal Learning Test, Rey-Osterrieth Complex Figure Test (RCFT) – recall and recognition), attention (Digit Span Test – forward and backward), executive function (Color Trails Test 1 and 2, Animal Fluency), language (modified Boston Naming Test), visuomotor speed (Symbol- Digit Modalities Test), and visuoconstruction (RCFT – copy). All raw scores were standardized against the NCI group at baseline to compute domain-specific and global cognitive z-scores for longitudinal analyses (23).

### APOE genotyping and plasma p-tau217 measurement

Ethylenediaminetetraacetic acid (EDTA) non-fasting blood was collected, pre-processed, and stored at −80°C at baseline following standard protocol. Genomic deoxyribonucleic acid (DNA) was extracted and purified using the QIAamp DNA Micro Kit and stored at 4°C until further processing. A DNA segment spanning the *APOE* polymorphisms was amplified using polymerase chain reaction (PCR), followed by restriction enzyme digestion using HhaI (13). The digested products were subsequently electrophoresed and visualized to determine the *APOE* genotypes.

Plasma was extracted, mixed well, aliquoted in 0.2 mL aliquots, and stored in Eppendorf tubes at −80°C until further processing (16). Plasma p-tau217 was measured on Single molecule array (Simoa) HD-X platform using the commercial ALZpath pTau217 assay (#104371, Quanterix Corp., Billerica,MA).

### Neuroimaging

*Image acquisition.* 3T brain MRI (MAGNETOM Trio™, Siemens, Germany) were conducted at baseline at the Centre for Translational Magnetic Resonance Research, National University of Singapore. High-resolution T1-weighted structural sequences were acquired using a magnetization-prepared rapid gradient echo sequence with 192 continuous sagittal slices (repetition time (TR) = 2300 ms, echo time (TE) = 1.9 ms, inversion time (TI) = 900 ms, flip angle = 9°, and field of view (FOV) = 256 × 256 mm^2^, voxel size = 1 × 1 × 1 mm^3^). T2 fluid-attenuated inversion recovery (FLAIR) sequence was acquired with 48 axial slices (TR = 9000 ms, TE = 82 ms, TI = 2500 ms, flip angle = 180^◦^, and FOV = 232 × 256 mm^2^, voxel size = 1 × 1 × 3 mm^3^).

### Image processing

Structural T1-weighted images were processed using an automated procedure in FreeSurfer 7.4 (https://surfer.nmr.mgh.harvard.edu/). The subcortical brain volumes were quantified by the subcortical segmentation pipeline (24) and regional cortical thickness were quantified by the cortical parcellation pipeline based on the Desikan-Killiany atlas (25). For each region-of-interest (ROI), bilateral measures were summed for subcortical volumes and averaged for cortical thickness (26). T2-FLAIR images were used to quantify total WMH volume using a deep convolutional network and ensemble models segmentation algorithm with high accuracy in clinical datasets (27, 28). Each participant’s WMH mask and T2-FLAIR scan were overlaid to visually inspect for any segmentation errors. Tract-based WMH were derived from the Johns Hopkins University white matter tractography atlas (29) using the spatially normalized WMH maps in FSL 6.0 (30).

### Statistical analysis

We performed separate Analysis of Covariance to examine if there were *APOE* ε4 (carriers vs. non-carriers) group differences in plasma p-tau217 concentration, subcortical volumes, cortical thickness, and WMH volumes. All models were adjusted for age, sex, years of education, and dementia diagnosis, with models involving cerebral volumetric measures further adjusted for total intracranial volume (TIV) to account for head size. Plasma p-tau217 levels and WMH volumes were log-transformed prior to model fitting due to right-skewed distributions. Interactions between *APOE* ε4 and diagnosis were additionally tested to determine if dementia had any influence on these associations. The *p*-values involving regional brain measures were further adjusted for multiple comparisons at a false discovery rate (FDR) of *q* < 0.05 for 8 subcortical regions, 34 cortical regions, and 11 white matter tracts.

Subsequently, we conducted serial mediation analysis (31, 32) to examine the mechanistic pathway through which *APOE* ε4 influences global and domain-specific cognitive decline. Among dementia-free participants (n = 396), a separate serial mediation model with survival outcome (31) was used to estimate hazards of incident dementia. In all mediation analyses, *APOE* ε4 served as the exposure (S), plasma p- tau217 as the upstream mediator (M_1_), and hippocampal volume or entorhinal thickness as the downstream mediator (M_2_). All biomarker measures were standardized prior to model fitting.

We decomposed the total effect of *APOE* ε4 into direct and indirect components, quantifying mediation through plasma p-tau217 alone, through hippocampal volume or entorhinal thickness alone, and through the serial pathway (S → M_1_ → M_2_ → outcome). Point estimates and 95% confidence intervals (CI) were obtained via Monte Carlo simulation (B = 5,000), and the proportion of the total effect mediated by each indirect pathway was computed. All models were adjusted for age, sex, education, and baseline diagnosis, with paths involving hippocampal volume further adjusted for TIV.

We performed sensitivity analyses including (i) adding total WMH volume as a covariate in the outcome models to account for vascular contributions, and (ii) stratifying by baseline diagnosis to assess whether the associations with global cognitive decline differed between patients with dementia and dementia-free participants. All analyses were performed using R Statistical Software (version 4.4.2; R Core Team 2024).

## Results

### Cohort characteristics

Cross-sectional analysis was conducted for up to 659 participants with available *APOE* and biomarker data (Figure 1). The excluded group of participants (n = 41) had fewer years of education (2 years, *p* = 0.015) and a higher rate of dementia (30.4%, *p* < 0.001). 194 participants (29.4%) were classified as *APOE* ε4 carriers, having at least one ε4 allele. The *APOE* genotypes (Figure 2A), in order of decreasing prevalence, were ε3/ε3 (60.2%), ε3/ε4 (24.7%), ε2/ε3 (9.1%), ε4/ε4 (3.2%), ε2/ε4 (1.5%), and ε2/ε2 (1.2%). *APOE* ε4 was more prevalent in CIND (χ^2^ = 13.35, *p* < 0.001) and AD dementia (χ^2^ = 14.98, *p* < 0.001), but not VAD (χ^2^ = 1.33, *p* = 0.249), compared to NCI (Figure 2B).

**Figure 1.**
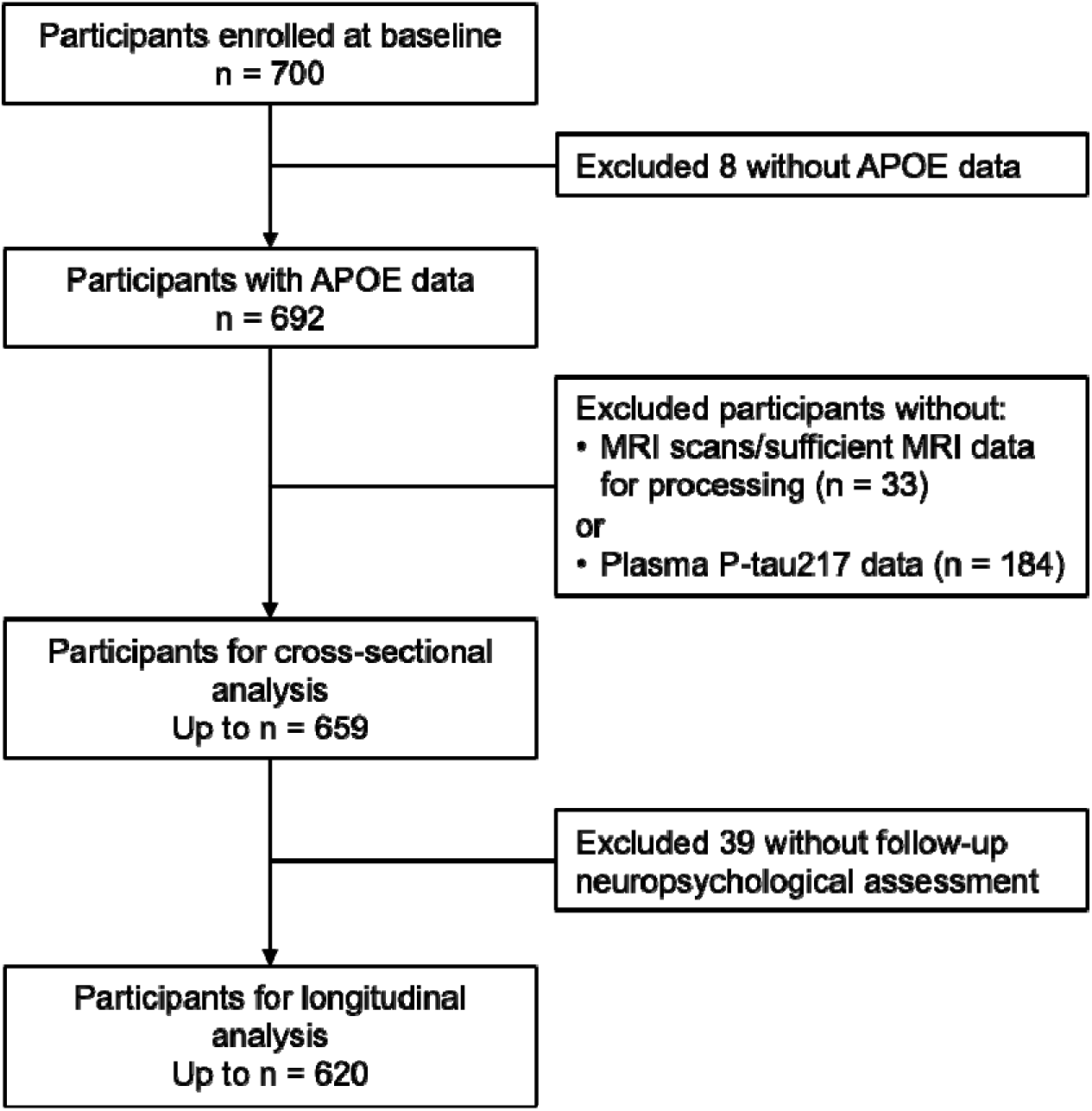
Participant flowchart.

**Figure 2.**
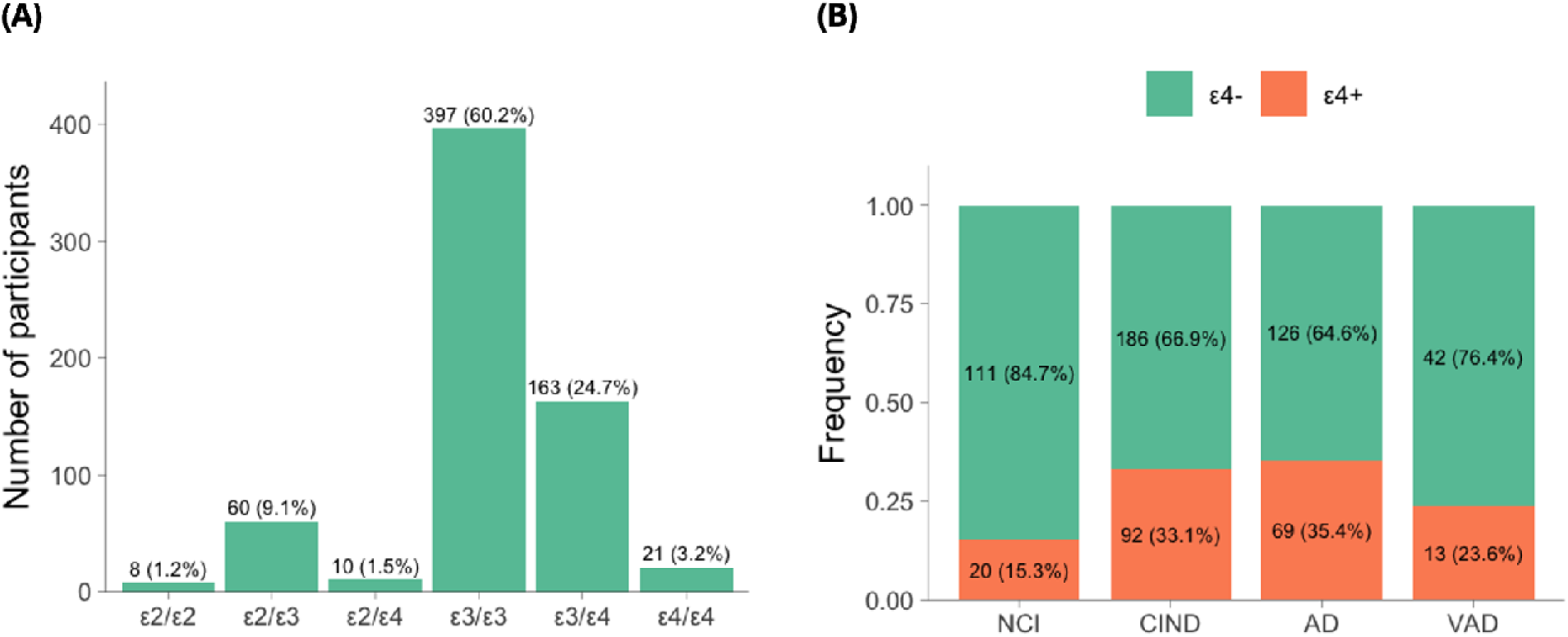
Prevalence of *APOE*. (A) shows the distribution of *APOE* genotypes and (B) shows the frequencies of *APOE* ε4 by diagnosis.

The *APOE* ε4 distribution did not differ significantly among CIND, AD dementia, and VAD (all *p* > 0.05). As summarized in Table 1, *APOE* ε4 carriers had a lower rate of hypertension (χ^2^ = 5.51, *p* = 0.019) but did not differ from non-carriers in other baseline characteristics.

**Table 1.** Baseline characteristics of study cohort.

| **Characteristics** | **All**<br>**n = 659** | ***APOE* ε4-**<br>**n = 465 (70.6%)** | ***APOE* ε4+**<br>**n = 194 (29.4%)** | ***p*** |
|---|---|---|---|---|
| Age, mean (SD) | 73.47 (8.01) | 73.50 (8.11) | 73.41 (7.79) | 0.906 |
| Female, n (%) | 372 (56.4) | 262 (56.3) | 110 (56.7) | 1.000 |
| Education, mean (SD) | 6.99 (5.06) | 6.94 (5.15) | 7.12 (4.85) | 0.668 |
| Ethnicity, Chinese, n (%) | 551 (83.6) | 397 (85.4) | 154 (79.4) | 0.075 |
| Hypertension, n (%) | 459 (69.7) | 337 (72.5) | 122 (62.9) | **0.019** |
| Hyperlipidemia, n (%) | 467 (70.9) | 337 (72.5) | 130 (67.0) | 0.189 |
| Diabetes mellitus, n (%) | 223 (33.8) | 163 (35.1) | 60 (30.9) | 0.352 |
| MMSE score, mean (SD) | 21.59 (6.07) | 21.79 (6.08) | 21.10 (6.03) | 0.186 |

Abbreviations: MMSE = Mini-Mental State Examination.

### APOE ε4 and differences in plasma p-tau217 and brain structures

Compared with non-carriers, *APOE* ε4 carriers had higher plasma p-tau217 levels (*F*_1,501_ = 74.58, *p* < 0.001) and smaller volumes of the hippocampus (*F*_1,649_ = 28.62, *p* < 0.001, *q* < 0.001), amygdala (*F*_1,649_ = 12.70, *p* < 0.001, *q* = 0.002), accumbens (*F*_1,649_ = 7.58, *p* = 0.006, *q* = 0.016), and putamen (*F*_1,649_ = 5.22, *p* = 0.023, *q* = 0.045) (Figure 3).

**Figure 3.**
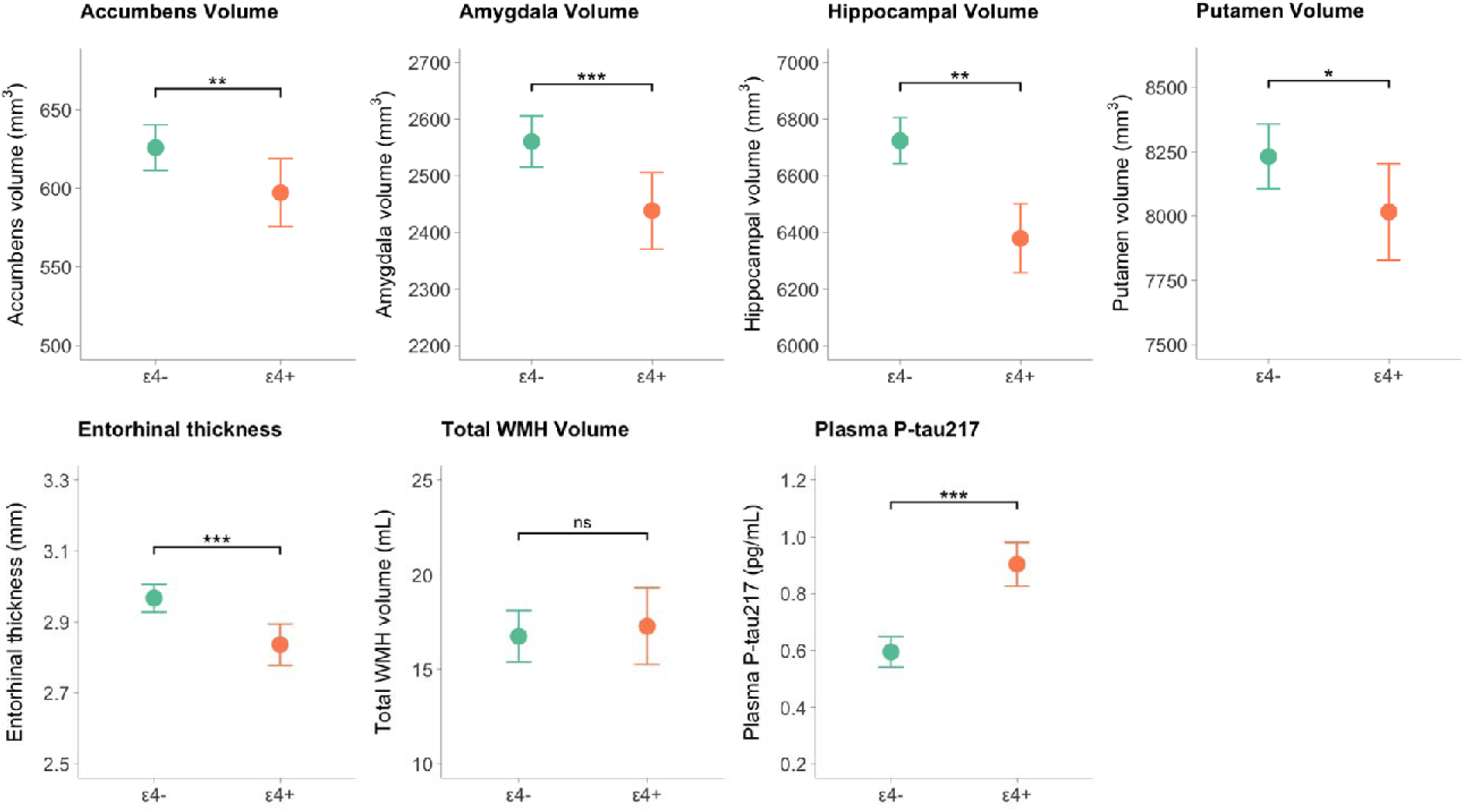
Adjusted *APOE* ε4 group differences in plasma p-tau217 and brain structures. Separate analyses of covariance were fitted for subcortical volumes, cortical thickness, plasma p-tau217 levels, and white matter hyperintensities (WMH) volumes, adjusted for age, sex, education, and dementia diagnosis. Models with cerebral volumetric measures were additionally adjusted for total intracranial volume. \**p* < 0.05, \*\**p* < 0.01, \*\*\**p* < 0.001.

The volumetric differences in the brainstem, caudate, pallidum, and thalamus were not statistically significant (all *p* > 0.05). Of the 34 cortical ROIs, *APOE* ε4 carriers had lower thickness in the entorhinal cortex (*F*_1,653_ = 17.98, *p* < 0.001, *q* = 0.001), isthmus cingulate (*F*_1,653_ = 6.44, *p* = 0.011), and lingual (*F*_1,653_ = 4.44, *p* = 0.035) but only entorhinal thickness remained statistically significant after adjustment for multiple comparisons. There were no statistically significant differences in global cortical thickness, total WMH volume, and tract-level WMH volumes. Dementia diagnosis also did not significantly influence these associations (*APOE* ε4 × dementia, all *p* > 0.05).

### Serial mediation by plasma p-tau217 and neurodegeneration on rate of cognitive decline

*APOE* ε4 was associated with faster decline in the cognitive domains of memory (*β* = - 0.035, 95% CI = −0.059, −0.011, *q* = 0.008), executive function (*β* = −0.022, 95% CI = - 0.043, −0.002, *q* = 0.035), language (*β* = −0.240, 95% CI = −0.407, −0.076, *q* = 0.008), and visuomotor speed (*β* = −0.024, 95% CI = −0.039, −0.009, *q* = 0.006), as well as global cognition (*β* = −0.065, 95% CI = −0.107, −0.024, *p* = 0.001). Of the total *APOE* ε4 effect on global cognitive decline (Figure 4), 70% was mediated through plasma p- tau217 (*β* = −0.046, 95% CI = −0.065, −0.029, *p* < 0.001), 13.8% through the serial pathway of plasma p-tau217 to hippocampal volume (*β* = −0.009, 95% CI = −0.015, - 0.004, *p* = 0.007), and 10% through hippocampal volume alone (*β* = −0.007, 95% CI = - 0.015, 0.000, *p* = 0.132). All indirect effects remained similar after accounting for total WMH volume (*β* = −0.027, 95% CI = −0.048, −0.006, *p* = 0.010). The direct effect of *APOE* ε4 on global cognitive decline was not significant (*β* = −0.004, 95% CI = −0.044, 0.036, *p* = 0.846).

**Figure 4.**
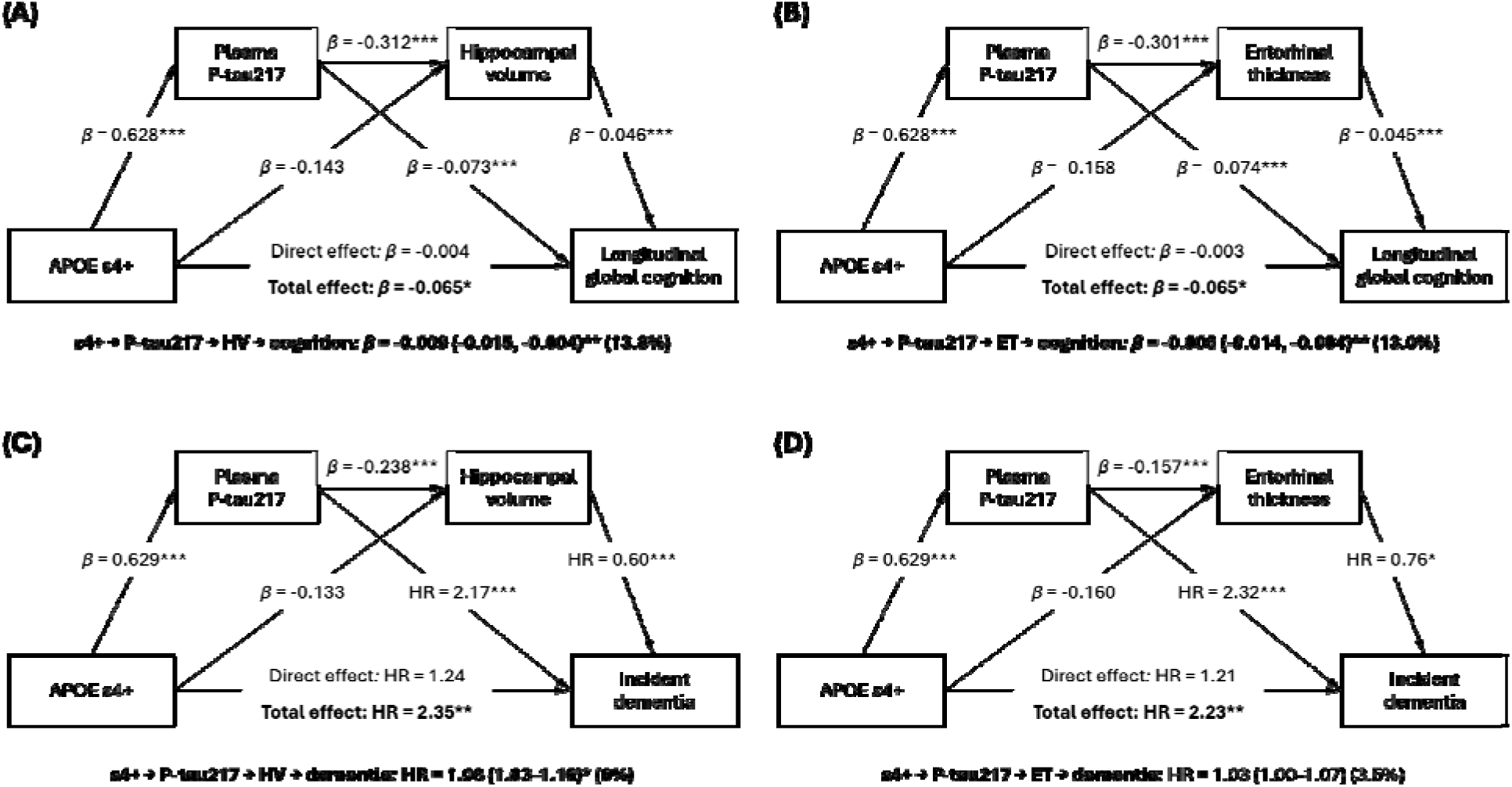
Serial mediation through plasma p-tau217 and neurodegeneration markers on clinical outcomes. Mediation analyses with linear regression and survival outcome showed the total, direct, and indirect effects of *APOE* ε4 on global cognitive decline and incident dementia hazard via serial pathways of plasma p-tau217 to hippocampal volume (A, C) and entorhinal thickness (B, D). Models were adjusted for age, sex, education, and dementia diagnosis. Paths involving hippocampal volume were additionally adjusted for total intracranial volume. \**p* < 0.05, \*\**p* < 0.01, \*\*\**p* < 0.001.

Similar proportions were mediated through the serial pathway of plasma p-tau217 to entorhinal thickness (13%, *p* = 0.006) and entorhinal thickness alone (10.9%, *p* = 0.120) (Figure 4), even after accounting for total WMH volume (*β* = −0.022, 95% CI = −0.041, - 0.003, *p* = 0.026). For domain-specific cognition, significant serial mediation through plasma p-tau217 to hippocampal volume and through plasma p-tau217 to entorhinal thickness were also observed for memory, attention, and visuomotor speed (q < 0.05, Table 2).

**Table 2.** Serial mediation of *APOE* ε4 on domain-specific cognitive decline through plasma p-tau217 and neurodegeneration markers.

|  | **Memory** | **Executive function** | **Attention** | **Language** | **Visuomotor speed** | **Visuoconstruction** |
|---|---|---|---|---|---|---|
| **Path** | **β (95%CI)** | **β (95%CI)** | **β (95%CI)** | **β (95%CI)** | **β (95%CI)** | **β (95%CI)** |
| **Model 1: HV as downstream mediator** |  |  |  |  |  |  |
| ε4 → PT → cognition | **-0.028 (-0.039, -0.018)\*‡** | **-0.011 (-0.019, -0.004)\*‡** | **-0.013 (-0.021, -0.005)\*‡** | **-0.193 (-0.273, -0.125)\*‡** | **-0.007 (-0.013, -0.002)\*‡** | **-0.017 (-0.029, -0.005)\*‡** |
| ε4 → HV → cognition | -0.004 (-0.010, 0.000) | -0.002 (-0.005, 0.000) | -0.003 (-0.007, 0.000) | -0.018 (-0.046, 0.001) | -0.002 (-0.005, 0.000) | -0.002 (-0.007, 0.001) |
| ε4 → PT → HV → cognition | **-0.006 (-0.009, -0.003)\*‡** | -0.003 (-0.005, 0.000) | **-0.004 (-0.007, -0.001)\*‡** | -0.024 (-0.047, -0.005)* | **-0.003 (-0.005, -0.001)\*‡** | -0.003 (-0.008, 0.001) |
| Total Indirect Effect | **-0.038 (-0.051, -0.026)\*‡** | **-0.015 (-0.024, -0.008)\*‡** | **-0.020 (-0.029, -0.011)\*‡** | **-0.236 (-0.321, -0.160)\*‡** | **-0.012 (-0.019, -0.006)\*‡** | **-0.023 (-0.036, -0.011)\*‡** |
| Direct Effect | 0.003 (-0.020, 0.026) | -0.007 (-0.027, 0.013) | 0.003 (-0.019, 0.025) | -0.004 (-0.166, 0.158) | -0.012 (-0.028, 0.004) | -0.010 (-0.043, 0.023) |
| Total Effect | **-0.035 (-0.059, -0.011)\*‡** | **-0.022 (-0.043, -0.002)\*‡** | -0.017 (-0.039, 0.005) | **-0.240 (-0.407, -0.076)\*‡** | **-0.024 (-0.039, -0.009)\*‡** | -0.033 (-0.066, -0.001)* |
| **Model 2: ET as downstream mediator** |  |  |  |  |  |  |
| ε4 → PT → cognition | **-0.030 (-0.042, -0.020)\*‡** | **-0.013 (-0.021, -0.006)\*‡** | **-0.013 (-0.022, -0.006)\*‡** | **-0.178 (-0.253, -0.113)\*‡** | **-0.009 (-0.015, -0.003)\*‡** | **-0.017 (-0.030, -0.006)\*‡** |
| ε4 → ET → cognition | -0.003 (-0.007, 0.000) | -0.001 (-0.003, 0.001) | -0.003 (-0.007, 0.000) | -0.031 (-0.071, 0.001) | -0.001 (-0.004, 0.000) | -0.003 (-0.008, 0.000) |
| ε4 → PT → ET → cognition | **-0.004 (-0.006, -0.001)\*‡** | -0.001 (-0.003, 0.001) | **-0.003 (-0.006, -0.001)\*‡** | **-0.038 (-0.062, -0.018)\*‡** | -0.002 (-0.003, 0.000) | -0.003 (-0.007, 0.000) |
| Total Indirect Effect | **-0.037 (-0.049, -0.025)\*‡** | **-0.015 (-0.023, -0.008)\*‡** | **-0.020 (-0.029, -0.011)\*‡** | **-0.247 (-0.336, -0.169)\*‡** | **-0.012 (-0.018, -0.006)\*‡** | **-0.024 (-0.037, -0.012)\*‡** |
| Direct Effect | 0.002 (-0.021, 0.025) | -0.007 (-0.028, 0.013) | 0.003 (-0.019, 0.025) | 0.007 (-0.153, 0.167) | -0.012 (-0.028, 0.003) | -0.009 (-0.042, 0.024) |
| Total Effect | **-0.035 (-0.058, -0.011)\*‡** | **-0.022 (-0.043, -0.002)\*‡** | -0.017 (-0.039, 0.005) | **-0.240 (-0.406, -0.076)\*‡** | **-0.024 (-0.039, -0.009)\*‡** | -0.033 (-0.065, 0.000)* |

All models were adjusted for age, sex, education, and dementia diagnosis. Paths involving hippocampal volume were additionally adjusted for total intracranial volume. Path-specific effects and 95% CIs were estimated via Monte Carlo simulation (B = 5000). \**p* < 0.05, ‡q < 0.05 (false discovery rate adjustment for 6 comparisons).
Abbreviations: ET = entorhinal thickness; HV = hippocampal volume; PT = plasma p-tau217.

In diagnosis-stratified analyses (Table S1), the mediation effects through plasma p- tau217 on global cognitive decline were stronger in dementia patients than dementia- free participants. However, a serial mediation through plasma p-tau217 to hippocampal volume was observed only in dementia-free participants while there was no evidence of serial mediation in dementia patients.

### Serial mediation by plasma p-tau217 and neurodegeneration on progression to dementia

A total of 79 (19.3%) dementia-free participants (5 NCI, 74 CIND) subsequently developed dementia. *APOE* ε4 was associated with a higher risk of incident dementia (HR = 2.35, 95% CI = 1.37, 4.06, *p* = 0.002). Of this effect, 58% was mediated through plasma p-tau217 (HR = 1.64, 95% CI = 1.33-2.13, *p* < 0.001), 9% through the serial pathway of plasma p-tau217 to hippocampal volume (HR = 1.08, 95% CI = 1.03-1.16, *p*= 0.028), and 8% through hippocampal volume alone (HR = 1.07, 95% CI = 0.97-1.22, *p*= 0.234). All indirect effects remained similar after accounting for total WMH volume (HR = 1.06, 95% CI = 0.78-1.41, *p* = 0.725). The direct effect of *APOE* ε4 on dementia risk was not significant (HR = 1.24, 95% CI = 0.75-2.04, *p* = 0.395). There was also no evidence of serial mediation through plasma p-tau217 to entorhinal thickness (3.5%, HR = 1.03, 95% CI = 1.00-1.07, *p* = 0.130) (Figure 4).

## Discussion

The current study shows, in our Asian cohort, that *APOE* ε4 was associated with higher plasma p-tau217 concentrations, reduced entorhinal thickness, and smaller volumes of the hippocampus, amygdala, accumbens, and putamen, but not with WMH burden. *APOE* ε4 was linked to faster cognitive decline and an increased risk of incident dementia, with these clinical effects primarily attributed to higher plasma p-tau217 and a sequential pathway from plasma p-tau217 to smaller hippocampal volume. The present results extend our previous cross-sectional findings (13) by demonstrating that *APOE* ε4 was associated with accumbens and putamen volumes, suggesting that its neurodegenerative impact may extend beyond medial limbic regions to implicate striatal pathways. The observed serial mediations of *APOE* ε4 through plasma p-tau217 and neurodegeneration, independent of WMH, suggest a biological mechanistic relationship whereby *APOE* ε4 contributes to poorer cognitive and clinical trajectories via AD-related pathology in older Asian adults.

Smaller samples from Japan and Taiwan have reported significant associations between *APOE* ε4 and reduced hippocampal volume, but evidence has been limited for its effects on other brain structures (33, 34). The hippocampus and amygdala, both associated with *APOE* ε4 in this study, are interconnected limbic regions that have been implicated in emotional dysregulation in Alzheimer’s and vascular diseases (35–37). With evidence in patients with AD that *APOE* ε4 may increase the risk of neuropsychiatric disturbances (38) and preclinical data linking the *APOE* ε4 allele and anxiety phenotype through pathological changes in the amygdala (39, 40), our findings point to a potential neuropsychiatric vulnerability arising from *APOE* ε4-related neurodegeneration of limbic structures.

Beyond these regions, *APOE* ε4 was also associated with reduced volumes of the accumbens and putamen. Both nuclei interact closely with limbic structures as key components of the striatum, supporting cognitive function, reward processing, and procedural learning (41), and have been linked to cognitive decline and dementia in Asian cohorts (42, 43). Together, our findings suggest increased susceptibility to *APOE* ε4-related subcortical limbic-striatal atrophy among older Asian adults, with the observed pattern of neurodegeneration largely aligning with current models of AD- related atrophy in which the hippocampal formation and entorhinal cortex are affected early, before cortical thinning becomes more widespread as the disease advances (44).

In addition to neurodegeneration, current AD staging frameworks emphasize amyloid-β and tau pathologies, and increasingly incorporate core fluid biomarkers, such as plasma p-tau217 (15). Owing to its close association with both cerebral amyloid-β and tau pathology, plasma p-tau217 is regarded as one of the most sensitive biomarkers for AD pathology (45). The dynamic between *APOE* ε4 and AD plasma biomarkers observed in European-ancestry cohorts is increasingly being replicated in Asian populations, where *APOE* ε4 carriers exhibited higher cerebral amyloid-β positivity, which was further associated with elevated plasma p-tau217 (16, 17). Clinical models further indicate that the effects of amyloid-β and tau biomarkers on cognition are partly mediated by brain atrophy, although the specific role of *APOE* ε4 was often not examined explicitly (46–48).

In this context, we observed substantial indirect effects of *APOE* ε4 on steeper rates of cognitive decline and higher risk of incident dementia via plasma p-tau217 and a serial pathway from p-tau217 to neurodegeneration, particularly hippocampal atrophy, in line with current concepts of *APOE* ε4-driven AD pathology (5). While both indirect pathways were statistically significant, most of the mediated effect was attributable to the plasma p-tau217-only pathway, suggesting that *APOE* ε4 influences cognitive trajectories primarily through amyloid-related tau dysregulation, with neurodegeneration representing a more downstream component or sequela. The distinctive biological properties of p-tau species including p-tau217, which reflects amyloid-β-related tau phosphorylation and release, hence capturing several phases of the amyloid-β cascade, may underlie these stronger mediating effects. Adding neurodegeneration as a downstream mediator did not substantially increase the explained pathway, especially in patients with dementia, likely because plasma p-tau217 and hippocampal volume both indexed overlapping pathological processes, such as medial temporal tau burden. Neurodegeneration may also introduce additional heterogeneity as it could be driven by multiple parallel mechanisms, including reactive microgliosis, neuroinflammation, and synaptic dysfunction (49, 50). Consistent with this mediation pattern, we found little evidence for direct effects of *APOE* ε4 on all outcomes, indicating that *APOE* ε4 status adds limited prognostic value once plasma p-tau217 is elevated, essentially capturing the downstream consequences of *APOE* ε4 on AD-related neurodegeneration.

Beyond the amyloid-induced mechanism, mediating effects of *APOE* ε4 through WMH burden and lobar microbleeds have also been reported in large European cohorts (11, 12), underscoring the contribution of vascular pathways to cognitive decline independent of amyloid-β or tau pathology. In our cohort, however, *APOE* ε4 was not significantly associated with WMH volume. Although there remains limited and inconsistent evidence in Asian populations to support a purely vascular pathway linking *APOE* ε4 to cognitive dysfunction (13, 14, 51), data from a Korean study indicate that vascular risk factors, particularly hypertension, may potentiate the effects of *APOE* ε4 on brain aging indices (9). Emerging clinical models on *APOE*-related alterations in lipid and glucose metabolism further suggest that alternative mechanisms contributing to *APOE*-related cognitive vulnerability require further investigation as potential targets alongside *APOE* polymorphism for risk assessment and intervention in Asian populations.

Although there is currently insufficient evidence to support population-wide *APOE* ε4 screening for AD (52), delineating *APOE* ε4 status may have valuable applications in clinical risk stratification and research. As amyloid-targeted therapies become more accessible, rapid and accurate *APOE* ε4 quantification (53), in combination with plasma p-tau217 profiling, may be essential for identifying individuals most likely to benefit from treatment and for stratifying risk of Amyloid-Related Imaging Abnormalities (54), thereby informing treatment selection and monitoring. Plasma p-tau217 may serve as a clinically actionable biomarker, especially in *APOE* ε4 carriers who may already exhibit early biological vulnerability, enabling risk stratification beyond genetic risk alone. Finally, *APOE* ε4 genotyping will continue to facilitate therapeutics development by enabling *APOE*-targeted strategies, such as modulating *APOE*-amyloid interactions, enhancing *APOE* lipidation, and regulating APOE receptor expression (3).

Among the strengths of this study are its longitudinal comprehensive neuropsychological assessments and the diverse cognitive profiles of the cohort, which better reflect real-world clinical settings. The integration of *APOE* ε4, plasma p-tau217, and detailed regional structural MRI measures also provided a unique opportunity to examine the succession from genetic risk to biomarker abnormalities and cognitive decline within a single analytic framework. However, several limitations warrant consideration. Although plasma p-tau217 is a sensitive biomarker of AD pathology, it remains an indirect indicator of cerebral amyloid-β burden, for which amyloid PET imaging would be required for definitive confirmation. Additionally, the effects of *APOE* ε4 on biomarker trajectories could not be inferred with only cross-sectional plasma p- tau217 and MRI data. Future studies incorporating longitudinal biomarker assessments, along with additional pathological processes, such as neuroinflammation and microstructural brain changes, will be important to more fully characterize the gene- pathophysiological dynamics.

In conclusion, this study identified an association between *APOE* ε4 and a pattern of subcortical vulnerability in older Asian adults, characterized by volumetric reductions in the amygdala, accumbens, hippocampus, and putamen, alongside entorhinal cortical thinning. Our findings suggest that *APOE* ε4 impairs cognition predominantly through an AD-specific pathway involving elevated plasma p-tau217 and medial temporal lobe atrophy, rather than through WMH burden. Collectively, these results provide population-specific evidence for a potentially tau-driven, regionally selective mechanism of *APOE* ε4 in Asians and support the value of peripheral tau biomarkers for risk stratification in this understudied population.

## Data Availability

All data produced in the present study are available upon reasonable request to the authors.

## Acknowledgements

None.

## Author Contributions

CNK (Conceptualization; Formal analysis; Data Curation; Writing - Original Draft); JRC (Data Curation; Writing - Review & Editing); DC (Data Curation); YLC (Data Curation; Writing - Review & Editing); NV (Writing - Review & Editing); BYT (Data Curation; Writing - Review & Editing); JHZ (Funding acquisition; Writing - Review & Editing); CLC (Data Curation; Funding acquisition; Writing - Review & Editing); MKPL (Data Curation; Writing - Review & Editing); CHT (Conceptualization; Supervision; Funding acquisition; Writing - Review & Editing).

## Statements and Declarations

### Ethics considerations

This study received ethics approval from the National Healthcare Group Domain- Specific Review Board (2018/01098).

### Consent to participate

Written informed consent was obtained before enrollment in the study.

### Consent for publication

Not applicable.

### Data Availability

The data supporting the findings of this study are available upon request. The data are not publicly available due to privacy or ethical restrictions.

